# Effectiveness of vaccination in preventing severe SARS CoV-2 infection in South India-a hospital-based cross-sectional study

**DOI:** 10.1101/2021.09.17.21263670

**Authors:** A Charles Pon Ruban, M Aazmi, K. Shantaraman

**Affiliations:** Department of Community Medicine, Tirunelveli Medical College, Tamil Nadu,India; Department of Pathology, Tirunelveli Medical College, Tamil Nadu, India

**Keywords:** Covid 19 vaccine, Effectiveness of Covid vaccine, SARS CoV-2, Severe covid 19

## Abstract

**Background & objectives:** Establishing concrete evidence on the effect of vaccination on the severity of SARS CoV-2 infections in real-world situations is the need of the hour. This study aims to estimate the effectiveness of Covid 19 vaccines in preventing the new and severe SARS CoV-2 infections.

**Design:** Cross-sectional study

**Setting& Participants:** We did this cross-sectional study among the 4565 patients consecutive adult inpatients admitted in the Covid 19 wards of a tertiary care hospital from May 7, 2021, to October 7, 2021, during the second wave of the Covid 19 pandemic. Information on basic demographic variables, RT PCR status, vaccination status, outcome and clinical severity of illness were obtained from the electronic hospital patient records.

**Results:** Only 4% of the study participants had prior vaccination. The type of vaccine and number of doses didn’t have any protective effect against the new SARS CoV-2 infection and breakthrough infection. Fully vaccinated RTPCR positive patients had an 82% reduction in the need for ICU admission (OR 0.09; AOR 0.18, CI (0.04 to 0.8), P <0.05) and a non-significant 79% in mortality (OR 0.19; AOR 0.21, CI (0.04 to 1.1) P>0.05).

**Conclusion:** Vaccination doesn’t protect against new SARS Cov-2 infection and breakthrough infection however significant protection was documented against severe SARS Cov-2 infection. The protective effect shown by the vaccines in preventing the severe form of SARS Cov-2 infection among fully vaccinated patients was 82%. Vaccination coverage should be increased urgently to halt the impending wave of severe acute respiratory syndrome coronavirus 2 (SARS-CoV-2) infection.

## Introduction

The impact caused by the Novel SARS-CoV-2 virus on global health is indescribable. As of October 4, 2021, globally 234,609,003 people were affected and 4,797,368 deaths were caused by SARS CoV-2. India too suffered badly and had documented 33,834,702 confirmed Covid 19 cases and 448,997 deaths[1]. Vaccination drive is one of the most imperative strategies ratified to combat the deadly pandemic. Eradication of the Covid19 may be too aspirational a goal given the current situation; elimination of Covid 19 is still a realistic goal. Israel has achieved an all-time low level of infections of SARS-CoV-2 with high vaccination coverage[2]. India being the pharmaceutical hub; there are 8 vaccines in the pipeline under various phases of the clinical trial and 2 vaccines – Covishield and Covaxin are in wide usage[3]. Covishield, produced by Serum Institute of India in synergy with Oxford University and Pharmaceutical giant Astra Zeneca uses a non-replicating adenovirus viral vector that infects chimpanzees given in 2 doses 84 days apart. The efficacy of the vaccine was found to be 76% after the first dose. If the interval between the first and second dose was extended to 12 weeks or more the efficacy rose to 91.6%[4]. Covaxin, a domestic vaccine developed in the country by Bharat Biotech in collaboration with the Indian Institute of Medical Research (ICMR) and subsidiary National Institute of Virology (assisted by isolating virus sample) is an inactivated vaccine given 28 days apart with a vaccine efficacy of 81%[5].

The national covid vaccination program was launched on 16 January 2021 in the country with the first phase covering Health Care workers and frontline workers. Considering high mortality in elderly and comorbid people, vaccination for greater than 60 years and with Comorbid diseases in 45-60 years age group was started from 1 March 2021. Further on all people in the age group of 45-60 years irrespective of the comorbid status were included in the vaccination drive from 1 April 2021. Vaccination for 18-44 years was kicked off from 1 May 2021[6].

Vaccine efficacy is defined as reduced risk of infection or disease among vaccinated individuals resulting from vaccination in carefully controlled circumstances; estimated from randomized clinical trials. Vaccine effectiveness is reduced risk of infection or disease among vaccinated individuals attributed to vaccination in real-world conditions; estimated from observational (non-randomized) studies. Gauging COVID-19 vaccine performance in the real world is crucial. Several factors like cold chain management during transportation and storage, completing dosing schedule, the inclusion of the general population (as people enrolled in a clinical trial are many times young and healthy) has a bearing on real-world vaccine effectiveness[7]. Vaccine hesitancy is one of the top ten global health threats reported by the World health organization which threatens the progress made against vaccine-preventable diseases[8]. In Tamil Nadu, the proportion of fully vaccinated people (18%) and the proportion of partially vaccinated people (51%) is still below the national average (fully vaccinated 20.4%, partially vaccinated 51.5%) though the vaccine coverage of the state has improved recently[9]. With high vaccine hesitancy among the population, providing concrete evidence on the effect of vaccination on the severity of the disease has become crucial and need of the hour. Hence this study was mapped out to analyse vaccine effectiveness in preventing severe SARS CoV-2 infection as there is a significant dearth of such studies in our country to arrive at a decisive policy on vaccination.

## Objective

1. To estimate the proportion of patients with severe SARS CoV-2 infections among the patients admitted in Covid 19 wards in a tertiary care hospital, South India
2. To estimate the occurrence of new SARS CoV-2 infections following Covid vaccination among the patients admitted in Covid 19 wards in a tertiary care hospital, South India.
3. To find out the effectiveness of Covid vaccination in preventing the severe SARS CoV-2 infections among the patients admitted in Covid 19 wards in a tertiary care hospital, South India

## Methodology

### Study setting

Triage Centre and Covid 19 ward of a tertiary care hospital, South India

### Study design

Cross-sectional study

### Study period

May 7, 2021, to October 7, 2021

### Study population

Inpatients admitted in Covid 19 wards

### Inclusion criteria

All adult patients (Aged ≥ 13 years) admitted in Covid 19 wards during the above said period were included.

### Exclusion criteria

Children (aged <13 years) were excluded from the study population as vaccination for them wasn’t started yet

Patients with incomplete data on outcome measured were excluded from the study.

### Sampling method

Consecutive sampling

### Sample size

NA

### Ethical committee approval

Institutional ethics committee approval was got. (Conducted on 19.08.2021)

### Study procedure

We did this cross-sectional study among the consecutive adult inpatients admitted in the Covid 19 wards of a tertiary care hospital, South India in the above said period. Our hospital is a government-run medical college hospital. It offers speciality and super speciality and allied health services and caters also to the neighbouring districts. All the patients admitted are tested for SARS-COV2 infection via nasopharyngeal and oropharyngeal swabs and the samples are transported to the designated Microbiological Lab within the College campus and the turnover time for report generation is within 12 hours.

An Electronic patient record database (Google sheet) has been developed and maintained by the hospital Covid control room and the required study variables were extracted from the database. After getting approval from the Institutional ethics committee, the following variables were collected; Name, age, sex, phone number, address, district, presence of symptoms, history of comorbid conditions, RT PCR status, SPO2, Respiratory rate, outcome, vaccination status, type of vaccine taken, number of doses, date of first positive and the difference between the date of RT PCR positive and date of vaccination. Self-reporting of vaccination status was confirmed through verification in the COWIN portal. The clinical severity of illness was categorized based upon the Respiratory Rate and Spo2 level measured at the time of admission.

Clinical guidelines for the management of Covid 19 issued by the Ministry of Health and family welfare, Government of India was followed for disease classification as follows:[10].

*Mild*: SpO2 >94 and RR<24/min; *Moderate*: SpO2 90 - 94 and RR 24 - 30 /min

*Severe:* SpO2 < 90 and RR >30 /min.

World health organization definition for SARI (severe acute respiratory illness infection) was used. SARI is defined as a hospitalized person with an acute respiratory infection, with a history of fever or measured fever of ≥ 38°C and cough with onset within the last 10 days[7].

Vaccine breakthrough infection is defined as the detection of SARS-CoV-2 RNA or antigen in a respiratory specimen collected from a person ≥14 days after they have completed all recommended doses of COVID-19 vaccine[11].

Vaccine effectiveness was calculated using the formula 1-AOR[7].

### Outcomes

Occurrence of Covid 19 infection following vaccination,

The proportion of patients with breakthrough infection.

The proportion of patients with severe Covid 19 (patients requiring oxygen; patients requiring ICU admission, the proportion of patients died) following vaccination.

The effectiveness of Covid 19 vaccines in preventing severe SARS-CoV-2 infection.

### Data analysis

Data entry was done with Microsoft Excel and data analysis was done with SPSS 23 Version. Frequencies and proportions were calculated as appropriate. Means, Range and standard deviation were calculated for quantitative variables. The difference between the proportions was calculated using the Chi-Square test. Differences between more than two proportions were calculated using the Chi-Square trends. Multi variate logistic regression will be done for adjusting the confounders.

## Results

A total of 5291 patients have been admitted in Covid 19 wards during the above said period. After excluding patients with incomplete data about the main outcome studied (absent Spo2; n=636) and children aged less than 12 years (=90) a total of 4565 patients were included in this study. The majority of the patients were male (56%). The majority of the patients belonged to the age group of 61-70 years (24.6%) followed by the 51-60 years (24.2%) age group. The mean age of the study population was 54.3 years and the standard deviation was 16.1; the Median was 56 years (Table 1).

**Table 1:** Distribution of Baseline variables and Vaccination status (n=4565)

|  |  |  |  |  |
| --- | --- | --- | --- | --- |
| Baseline Variables | Vaccination status |  |  | P value |
|  | YES(n=187) | NO(N=4378) | Odd's ratio (CI) |  |
| Sex wise distribution |  |  |  |  |
| Male | 111(4.3%) | 2452(95.7%) | 1.2<br>(0.8 to 1.5) | 0.4 |
| Female | 76(3.8%) | 1926(96.2%) |  |  |
| Age-wise distribution |  |  |  |  |
| 13 to 18 years | 0 | 42(100%) | Chi square for trend<br>P =0.09 |  |
| 19 to 30 years | 27(6.1%) | 417(93.9%) |  |  |
| 31 to 40 years | 18(3.8%) | 457(96.2%) |  |  |
| 41 to 50 years | 28(3.8%) | 688(96.2%) |  |  |
| 51 to 60 years | 50(4.5%) | 1053(95.5%) |  |  |
| 61 to 70 years | 38(3.4%) | 1086(96.6%) |  |  |
| 71 to 80 years | 21(4%) | 494(96%) |  |  |
| 81 to 90 years | 4(2.9%) | 132(97.1%) |  |  |
| 91 to 100 years | 0 | 8(100%) |  |  |
| Presence of co-morbid conditions |  |  |  |  |
| Yes | 92(4%) | 2180(96%) | 1<br>(0.7 to 1.3) | 0.8 |
| No | 95(4.1%) | 2198(95.9%) |  |  |
| Clinical Category wise distribution |  |  |  |  |
| Mild | 103(7.4%) | 1293(92.6%) |  |  |
| Moderate | 25(3.9%) | 615(96.1%) | 0.5 | 0.003 |
| Severe | 59(2.3%) | 2470(97.7%) | 0.3 | 0 |

\*p value <0.05

84% of the patients were symptomatic and 50 % of the patients had co-morbid conditions. Among the patients presented with SARI (Severe acute respiratory illness), 44% were RT-PCR positive. Only 4% of the study population had received a covid 19 vaccine. Most of the cases were of mild type (Table 1).

Our study had found that 70% of patients with SARI and 66 % of the RT PCR positive patients required oxygen. 55% of the SARI patients and 50% of the RT PCR positive patients needed ICU admission. 20 % of the SARI patients and 25% of RTPCR positive patients had died during the hospital stay.

### Effectiveness of Covid 19 vaccine in preventing new SARS CoV 2 infection

2027 patients were RTPCR positive. Our study didn’t find any protective effect of vaccination against new COVID 19 infections. Type of vaccine nor number of doses has any effect on new infection. Females (1.2 times), people vaccinated with at least one dose of Covid 19 vaccine (1.8 times) and people who had 2 doses (2 times) and people who had Covaxin (2.3 times) had higher odds of becoming infected with SARS CoV 2 virus and they were statistically significant also (p<0.05). However, after multiple logistic regression analyses, none of them was found to be statistically significant (Table 2).

**Table 2:** Effectiveness of Covid 19 vaccine in preventing new/breakthrough SARS CoV 2 infection

| I. Occurrence of new SARS CoV 2 infection (n=4565) |  |  |  |  |  |
| --- | --- | --- | --- | --- | --- |
|  | Category | RT PCR Positive (n=2027) | RT PCR Negative (n=2538) | Odd's ratio (95%CI) | Adjusted OR (95%CI) |
| Sex | Male | 1092(42.6%) | 1471(57.4%) | 0.9<br>(0.8 to 0.9) * | 0.6<br>(0.3 to 1.2) |
|  | Female | 935(46.7%) | 1067(53.3%) |  |  |
| Age >56 years | Yes | 996(43.9%) | 1273(56.1%) | 0.96<br>(0.8 to 1) | 0.7<br>(0.4 to 1.3) |
|  | No | 1031(44.9%) | 1265(55.1%) |  |  |
| Presence of co-morbid conditions | Yes | 1012(44.5%) | 1260(55.5%) | 1<br>(0.9 to 1.1) | 1<br>(0.6 to 1.9) |
|  | No | 1015(44.3%) | 1278(55.7%) |  |  |
| Vaccinated with at least one dose | Yes | 108(57.8%) | 79(42.6%) | 1.8*<br>(1.3 to 2.4) * |  |
|  | No | 1919(43.8%) | 2459(56.2%) |  |  |
| Number of doses | 0 dose (n=4378) | 1919(43.8%) | 2459(56.2%) |  |  |
|  | 1 dose (n=135) | 76(56.3%) | 59(43.7%) | 1.6* |  |
|  | 2 doses (n=52) | 32(61.5%) | 20(38.5%) | 2* | 1.3<br>(0.6 to 2.4) |
| Type of vaccine | 0 dose (n=4378) | 1919(43.8%) | 2459(56.2%) |  |  |
|  | Covishield (n=145) | 81(55.9%) | 64(44.1%) | 1.6* |  |
|  | Covaxin (n=42) | 27(64.3%) | 15(35.7%) | 2.3* | 1.4<br>(0.7 to 2.9) |
| II. Breakthrough infection (n=45) |  |  |  |  |  |
|  | Category | Yes (n=23) | No (n=22) | Odd's ratio (95%CI) | Adjusted OR (95%CI) |
| Sex | Female | 10(58.8%) | 7(41.2%) | 1.6<br>(0.5 to 5.6) | 1.6<br>(0.4 to 6) |
|  | Male | 13(46.4%) | 15(53.6%) |  |  |
| Age >56 years | Yes | 9(42.9%) | 12(57.1%) | 0.5<br>(0.2 to 1.8) | 0.33<br>(0.1 to 1.5) |
|  | No | 14(58.3%) | 10(41.7%) |  |  |
| Presence of co-morbid conditions | Yes | 9(47.4%) | 10(52.6%) | 0.8<br>(0.2 to 2.5) | 1.2<br>(0.3 to 5.1) |
|  | No | 14(53.9%) | 12(46.1%) |  |  |
| Type of vaccine | Covaxin(n=15) | 11(73.3%) | 4(26.7%) | 4.1*<br>( 1 to 16) | 5.7*<br>(1.3 to 26) |
|  | Covishield(n=30) | 12(40%) | 18(60%) |  |  |

\*p value <0.05

### Effectiveness of Covid 19 vaccine in preventing breakthrough SARS CoV 2 infection

Out of the 52 patients who had 2 doses of vaccines, the data on days between the second dose of vaccination and date of RT PCR positive was available only for 45 patients. 23 of them (51%) had a breakthrough infection (RT PCR positive after 14 days of vaccination). Sex, age, presence of comorbid status and type of vaccine were considered for cross-tabulation. People vaccinated with Covaxin (OR 4.1 times, AOR 5.7(95% CI 1.3 to 26)) had higher odds of becoming infected with the SARS CoV 2 virus after 14 days of vaccination and it was statistically significant also (p<0.05) (Table 2)

### Effectiveness of Covid 19 vaccine in preventing a severe form of illness among SARI patients

4565 patients were admitted with the features of Severe Acute Respiratory infection (SARI). Males (1.2 times), those aged more than 56 years (2.1 times) and patients with co-morbidity (1.4 times) have higher odds of requiring ICU care and they were statistically significant(p<0.05). Males (1.3 times), those aged more than 56 years (2 times) and patients with co-morbidity (2.6 times) have higher odds of dying and they were statistically significant(p<0.05).

All the above factors were considered for multivariate logistic regression. The risk was augmented for baseline variables. Males (AOR 2.4, 95% CI (1.2 to 5)), those aged more than 56 years (AOR 2.5, 95% CI (1.2 to 5)) and patients with co-morbidity (AOR 2.9, 95% CI (1.4 to 5.9)) have higher odds of requiring ICU care and they were statistically significant(p<0.05). Males (AOR 4, 95% CI (1.3 to 12.8) and patients with co-morbidity (AOR 6.3, 95% CI (1.9 to 20.3) have higher odds of dying and they were statistically significant(p<0.05).

Partially vaccinated patients admitted with Severe acute respiratory infection (SARI) had a 55% reduction in the need for ICU admission and 30% fewer odds of dying. Fully vaccinated SARI patients had an 84% reduction in the need for ICU admission and 76% fewer odds of dying; both of them were statistically significant(p<0.05). However, after logistic regression analysis, fully vaccinated SARI patients had a 71% significant reduction (AOR 0.29, 95% CI (0.1 to 0.7)) in the need for ICU admission but the reduction shown in mortality had become non-significant. The covaxin vaccine offered significantly higher protection against the severe form of illness as compared to the Covishield vaccine. Patients with Covishield vaccination had 60% less ICU admission need and 40 % less risk of death whereas those vaccinated with Covaxin had an 86% reduction in the need for ICU admission and 60% fewer deaths. It was statistically significant(p<0.05). However, after logistic regression analysis, the adjusted odds ratio (AOR) for the reduction in ICU admission of Covaxin was 0.63(95% CI 0.3 to 1.5) and for mortality reduction, AOR was 0.75 (95% CI 0.2 to 2.5); both of them were not significant (Table 3).

**Table 3:** Effectiveness of Covid 19 vaccine in preventing a severe form of illness among SARI patients

| Baseline variables VS Outcome (n=4565) |  |  |  |  |  |  |  |  |  |
| --- | --- | --- | --- | --- | --- | --- | --- | --- | --- |
|  |  | Patients requiring ICU admission |  |  |  | Death |  |  |  |
| Variables | Category | Yes<br>(n=2529) | No<br>(n=2036) | Odd's<br>ratio<br>(95%CI) | Adjusted<br>OR<br>(95%CI) | Yes<br>(n=912) | No<br>(n=3653) | Odd's<br>ratio<br>(95%CI) | Adjusted OR<br>(95%CI ) |
| Sex | Male | 1478<br>(57.7%) | 1085<br>(42.3%) | 1.2<br>(1.1 to 1.4) * | 2.4<br>(1.2 to5)<br>* | 558<br>(21.8%) | 2005<br>(78.2%) | 1.3<br>(1.1 to 1.5) * | 4<br>(1.3 to 12.8) * |
|  | Female | 1051<br>(52.5%) | 951<br>(47.5%) |  |  | 354<br>(17.7%) | 1648<br>(82.3%) |  |  |
| Age >56<br>years | Yes | 1463<br>(64.5%) | 806<br>(35.5%) | 2.1<br>(1.9 to 2.4) * | 2.5<br>(1.2 to 5)<br>* | 576<br>(25.4%) | 1693<br>(74.6%) | 2<br>(1.7 to 2.3) * | 1<br>(0.4 to 2.7) * |
|  | No | 1066<br>(46.4%) | 1230<br>(53.6%) |  |  | 336<br>(14.6%) | 1960<br>(85.4%) |  |  |
| Presence of<br>co-morbid<br>conditions | Yes | 1344<br>(59.2%) | 928<br>(40.8%) | 1.4<br>(1.2 to 1.5) * | 2.9<br>(1.4 to 5.9) * | 622<br>(27.4%) | 1650<br>(72.6%) | 2.6<br>(2.2 to 3)<br>* | 6.3<br>(1.9 to 20.3) * |
|  | No | 1185<br>(51.7%) | 1108(43.3<br>%) |  |  | 290<br>(12.7%) | 2003<br>(87.4%) |  |  |
| Vaccination status Vs Outcome |  |  |  |  |  |  |  |  |  |
| Vaccinated<br>with at<br>least one<br>dose | Yes | 59<br>(31.6%) | 128<br>(68.5%) | 0.4<br>(0.3 to 0.5) * |  | 24<br>(12.8%) | 163<br>(87.2%) | 0.6<br>(0.4 to 0.9) * |  |
|  | No | 2470<br>(56.4%) | 1908<br>(43.6%) |  |  | 888<br>(20.3%) | 3490<br>(79.7%) |  |  |
| Number of<br>doses | 0 dose<br>(n=4378) | 2470<br>(56.4%) | 1908<br>(43.6%) |  |  | 888<br>(20.3%) | 3490<br>(79.7%) |  |  |
|  | 1 dose<br>(n=135) | 50<br>(37%) | 85<br>(63%) | 0.45* | 0.29<br>(0.1 to 0.7) | 21<br>(15.6%) | 114<br>(84.4%) | 0.7 | 0.29<br>(0.1 to 1.1) |
|  | 2 doses<br>(n=52) | 9<br>(17.3%) | 43<br>(82.7%) | 0.16* |  | 3<br>(5.8%) | 49<br>(94.2%) | 0.24* |  |
| Type of<br>vaccine | No<br>vaccinatio<br>n<br>(n=4378) | 2470<br>(56.4%) | 1908<br>(43.6%) |  |  | 888<br>(20.3%) | 3490<br>(79.7%) |  |  |
|  | Covishiel<br>d<br>(n=145) | 49<br>(33.8%) | 96<br>(66.2%) | 0.4* | 0.63<br>(0.3 to 1.5) | 20<br>(13.8%) | 125<br>(86.2%) | 0.6 | 0.75<br>(0.2 to 2.5) |
|  | Covaxin<br>(n=42) | 10<br>(23.8%) | 32<br>(76.2%) | 0.24* |  | 4<br>(9.5%) | 38<br>(90.5%) | 0.4* |  |

\*p value <0.05

### Effectiveness of Covid 19 vaccine against severe SARS CoV-2 infection

Males (1.3 times), those aged more than 56 years (2 times) and patients with co-morbidity (1.6 times) have higher odds of requiring ICU care. Males (1.3 times), those aged more than 56 years (2.2 times) and patients with co-morbidity (3 times) have higher odds of dying and they were statistically significant(p<0.05).

All the above factors were considered for multivariate logistic regression. The risk was augmented significantly for the baseline variables. Males had only higher odds of ICU admission need (AOR 2.9 (95% CI 1 to 8.8)) not for mortality. Patients with co-morbidity had significantly higher odds of ICU admission need (AOR 5.8 (95% CI 1.8 to 18.6) and mortality risks (AOR 10.3(95% CI (2 to 52.6)).

Partially vaccinated RTPCR positive patients had a 60% reduction in the need for ICU admission and 30% fewer odds of dying. Fully vaccinated RTPCR positive patients had a 91% reduction in the need for ICU admission and 81% fewer odds of dying; both were statistically significant(p<0.05). However, after logistic regression analysis, fully vaccinated RTPCR positive patients had an 82% significant reduction (AOR 0.18, 95% CI (0.04 to 0.8)) in the need for ICU admission but the reduction shown in mortality (AOR 0.21, 95% CI (0.04 to 1.1)) had become non-significant. The protective effect shown by the vaccines in preventing a severe form of SARS Cov-2 infection among fully vaccinated patients was 82%.

The covaxin vaccine offered significantly higher protection against severe SARS Cov-2 infection as compared to the Covishield vaccine. Patients with Covishield vaccination had 67% less ICU admission need and 40 % less risk of death whereas those vaccinated with Covaxin had an 85% reduction in the need for ICU admission and 60% fewer deaths. It was statistically significant(p<0.05). However, after logistic regression analysis, the adjusted odds ratio (AOR) for the reduction in ICU admission of Covaxin was 0.85(95% CI 0.2 to 3.3) and for mortality, AOR was 1.3 (95% CI 0.3 to 6.1); both of them became not significant (Table 4).

**Table 4:** Effectiveness of Covid 19 vaccine against severe SARS CoV-2 infection

|  |  |  |  |  |  |  |  |  |  |
| --- | --- | --- | --- | --- | --- | --- | --- | --- | --- |
| I. Baseline variables Vs Outcome (n=2027) |  |  |  |  |  |  |  |  |  |
|  |  | Patients requiring ICU admission |  |  |  | Death |  |  |  |
| Variables |  | Yes<br>(n=1050) | No<br>(n=977) | Odd's<br>ratio<br>(95%CI) | Adjusted<br>OR<br>(95%CI) | Yes<br>(n=511) | No<br>(n=1516) | Odd's<br>ratio<br>(95%CI) | Adjusted<br>OR<br>(95%CI) |
| Sex | Male | 601<br>(55%) | 491<br>(45%) | 1.3<br>(1.1 to<br>1.6) * | 2.9<br>(1 to<br>8.8) * | 299<br>(27.4%) | 793<br>(72.6%) | 1.3<br>(1.1 to<br>1.6) * | 3.3<br>(0.9 to<br>12.5) |
|  | Female | 449<br>(48%) | 486<br>(52%) |  |  | 212<br>(22.7%) | 723<br>(77.3%) |  |  |
| Age >56<br>years | Yes | 611<br>(61.4%) | 385<br>(38.6%) | 2<br>(1.8 to<br>2.6) * | 2.2<br>(0.8 to<br>6.4) | 325<br>(32.6%) | 671<br>(67.4%) | 2.2<br>(1.8 to<br>2.7) * | 1.3<br>(0.4 to<br>4.5) |
|  | No | 439<br>(42.6%) | 592<br>(57.4%) |  |  | 186<br>(18%) | 845<br>(82%) |  |  |
| Presence<br>of co-morbid<br>conditions | Yes | 580<br>(57.3%) | 432<br>(42.7%) | 1.6(1.3<br>to 1.9) * | 5.8<br>(1.8 to<br>18.6) * | 358<br>(35.4%) | 654<br>(64.6%) | 3<br>(2.5 to<br>3.8) * | 10.3<br>(2 to<br>52.6) * |
|  | No | 470<br>(46.3%) | 545<br>(53.7%) |  |  | 153<br>(15%) | 862<br>(85%) |  |  |
| II. Vaccination status Vs Outcome |  |  |  |  |  |  |  |  |  |
| Vaccinated | Yes | 26<br>(24%) | 82(76%) | 0.3<br>(0.2 to<br>0.4) * |  | 17<br>(15.7%) | 91<br>(84.3%) | 0.5<br>(0.3 to<br>0.9) * |  |
|  | No | 1024<br>(53.4%) | 895<br>(46.6%) |  |  | 494<br>(25.7%) | 1425<br>(74.3%) |  |  |
| Number of<br>doses | 0 dose<br>(n=4378) | 1024<br>(53.4%) | 895<br>(46.6%) |  |  | 494<br>(25.7%) | 1425<br>(74.3%) |  | 0.21<br>(0.04 to<br>1.1) |
|  | 1 dose<br>(n=135) | 23<br>(30.3%) | 53<br>(69.7%) | 0.4* | 0.18<br>(0.04 to<br>0.8) * | 15<br>(19.7%) | 61<br>(80.3%) | 0.7 |  |
|  | 2 doses<br>(n=52) | 3<br>(9.4%) | 29<br>(90.6%) | 0.09* |  | 2(6.3%) | 30<br>(93.7%) | 0.19* |  |
| Type of<br>vaccine | No<br>vaccination<br>(n=4378) | 1024<br>(53.4%) | 895<br>(46.6%) |  |  | 494<br>(25.7%) | 1425<br>(74.3%) |  | 1.3<br>(0.3 to<br>6.1) |
|  | Covishield<br>(n=145) | 22<br>(27.2%) | 59<br>(72.8%) | 0.33* | 0.85<br>(0.2 to<br>3.3) | 14<br>(17.3%) | 67<br>(82.7%) | 0.6 |  |
|  | Covaxin<br>(n=42) | 4(14.8%) | 23(85.2%) | 0.15* |  | 3(11.1%) | 24(88.9%) | 0.36* |  |

\*p value <0.05

## Discussion

We did this cross-sectional study among the patients admitted in the Covid 19 wards of a tertiary care hospital during the second wave of the Covid 19 pandemic. In our study, only 4% of SARI patients and 5.3 % of the RTPCR positive patients were vaccinated which is comparable to the district data as well as the state vaccine coverage during the initial phase. But it didn’t reflect the current vaccine coverage. There is a possibility that vaccination could have reduced the need for hospitalization.

Our study couldn’t find any protective effect of vaccination against new COVID 19 infections. Type of vaccine nor number of doses of vaccines has any effect on the occurrence of infection. 23 of them (51%) had a breakthrough infection. People vaccinated with Covaxin (OR 4.1 times, AOR 5.7(95% CI 1.3 to 26)) had got a breakthrough infection as compared to the Covishield vaccine. This is in contrast to the findings from other studies. Victor et al had documented that the protective effect of covid vaccination in preventing infection, hospitalization, need for oxygen and ICU admission were 65%, 77%, 92% and 94% respectively[12]. VIN-WIN cohort study conducted among 1.59 million health care workers and frontline workers of Indian Armed Forces showed a 93% reduction in breakthrough infection following Covishield vaccination[13]. Ella et al[5], Voysey et al[4] had also documented the protective effect of the vaccine in preventing infection. Pooled analysis of randomized control trials on ChAdOx1 nCoV-19 vaccines by Voysey et al showed that two standard doses had given 66.7% protection against hospitalization and symptomatic infections[14]. The efficacy of the covishield vaccine against symptomatic infections was found to be 76% after the first dose. If the interval between the first and second dose was extended to 12 weeks or more the efficacy rose to 91.6%[4]. But we couldn’t find any protective effect of the vaccine in preventing the infection following vaccination. Less number of vaccinated patients and lesser hospitalizations among them in our study might be one of the reasons. Study design used in this study also might be a factor. The difference in health-seeking behaviour among the vaccinated and non-vaccinated patients could have resulted in different rates of hospital admission also[7].

The proportion of SARI patients and RT PCR positive patients who required oxygen support, ICU admission and died reported by our study was significantly higher than the first wave studies and also other studies done during the second wave in India. Our study had found that 70% of patients with SARI and 66 % of the RT PCR positive patients required oxygen. 55% of the SARI patients and 50% of the RT PCR positive patients needed ICU admission. 20 % of the SARI patients and 25% of RTPCR positive patients had died during the hospital stay. Kumar G et al had reported that 50 % of patients needed oxygen support and high mortality during the second wave which is in line with our findings. This is in contrast to the finding that only 5-15% percentage of the patients had the severe disease as reported by the first wave study findings[15–20]. More transmission and more virulence were noticed in the second wave which suggests that this could be due to different virus strains[21]. Another important reason for the high proportion of patients with severe disease could be the hospital admission policy.

In our study, it was well documented that vaccination offered significant protection against severe SARS Cov-2 infection. The protective effect shown by the vaccines in preventing a severe form of SARS Cov-2 infection among fully vaccinated patients was 82%. Fully vaccinated SARI patients had a significant 84% (OR 0.16; AOR 0.29, 95% CI (0.1 to 0.7)) reduction in the need for ICU admission and a non-significant 76% (OR 0.24; AOR 0.29, 95% CI (0.1 to 1.1)) fewer odds of dying. Fully vaccinated RTPCR positive patients had a statistically significant 91% reduction in the need for ICU admission and 81% fewer odds of dying; However, after logistic regression analysis, fully vaccinated RTPCR positive patients had an 82% significant reduction (AOR 0.18, 95% CI (0.04 to 0.8)) in the need for ICU admission but the reduction shown in mortality (AOR 0.21, 95% CI (0.04 to 1.1)) had become non-significant.

Covaxin vaccine seemed to be offering significantly higher protection against severe SARS Cov-2 infection as compared to the Covishield vaccine before logistic regression. However, after logistic regression analysis, both of them became not significant. The adjusted odds ratio (AOR) for the reduction in ICU admission for Covaxin was 0.85(95% CI 0.2 to 3.3) and for mortality, AOR was 1.3 (95% CI 0.3 to 6.1).

The protective effect shown by the vaccines in preventing a severe form of the disease found in our study is comparable with other studies except for mortality. Less number of patients with vaccination and fewer number deaths among the vaccinated people reported in our study could be the reason for this. More samples are needed to conclude further. Victor et al have reported a high protective effect of vaccination in preventing the need for oxygen (92%) and ICU admission (94%) among health care workers in India[12]. Jaiswal et al had documented 95 % effectiveness in preventing death among fully vaccinated people(High-risk group)[22]. Christie et al have documented that improved vaccination coverage has reduced hospital admissions, the need for oxygen supplementation and ICU admission in the United States of America[23]. After vaccinating more than 70% of the population with two doses of BNT162b2, Israel had significantly reduced hospitalization and ICU admission[24]. Bernel et al have reported the real-world effectiveness of oxford Astra Zeneca in preventing ICU admissions as 73% in England[25]. Phase 3 trial results revealed that Covaxin had a protective effect of 93.4% against severe infections[26].The Vin-Win cohort study documented that the Covishield vaccine had shown 93% effectiveness against severe illness[13].

This is one of the unique studies in that effectiveness of covid vaccination among the SARI cases was also documented. The inclusion of a greater number of vaccinated people may give more evidence. There is a paucity of studies in this field and more studies with bigger sample sizes are needed to conclude further.

### Limitation

The proportion of vaccinated and fully vaccinated patients, deaths among vaccinated people were low in our study so the results should be interpreted cautiously. Explicit research with more sample size and negative case-control study is quintessential to corroborate the findings.

## Conclusion

Our study has found that vaccination didn’t protect against new SARS Cov-2 infection. The proportion of SARI patients and RT PCR positive patients who required oxygen support, ICU admission and died was significantly high during the second wave. Covid vaccination significantly reduced the severity of illness among SARI patients and even more reduction in risk was documented among the RT PCR positive patients. The protective effect shown by the vaccines in preventing a severe form of SARS Cov-2 infection among fully vaccinated patients was 82%. Vaccination coverage should be increased urgently to halt the impending wave of severe acute respiratory syndrome coronavirus 2 (SARS-CoV-2) infection

## Data Availability

https://docs.google.com/spreadsheets/d/1BLMMPTZz4MfYBX9dhP0Za6_8I_Gm__c__5nhkYf-ggQ/edit?usp=sharing
(ORIGINAL DATA IS AVAILABLE IN THE ABOVE LINK)

